# Estimation of infection rate and the population size potentially exposed to SARS-CoV-2 in Japan during 2020

**DOI:** 10.1101/2021.02.01.21250971

**Authors:** Motohiko Naito

## Abstract

**Background:** The infectious respiratory disease COVID-19, caused novel severe acute respiratory syndrome coronavirus 2 (SARS-CoV-2) reached pandemic status during 2020. The primary statistic data are important to survey the actual circumstances of COVID-19. Here, we report the analysis of the primary data of COVID-19 in Japan during 2020.

**Methods:** Data were collected and released systematically under Japan domestic law. Machine learning was conducted to estimate the positive rate in Japan and four prefectures (Tokyo, Osaka, Chiba, and Fukuoka).

**Results:** Primary data analysis revealed there were at least two peaks of infection in Japan; the first one was during April 2020 and the second one started from November 1, 2020. Estimating the positive rate in Japan as well as in the four prefectures reinforced the above observations. The positive rate in Japan during 2020 was estimated to be around 6% to 8%. We also estimated that 1.95 million people were possibly exposed to the novel virus on October 31, 2020. The numbers of related deaths were over 3,000 people at the end of 2020.

**Conclusion:** We estimated the infection rate of SARS-CoV-2 in Japan to be 6–8% in 2020. We also concluded that Japan had at least two infection-spreading periods, the first one being from Jan 19, 2020 until May 2020, and the second one beginning from November 1, 2020. Importantly, our analysis supports the need for clear definition of the criteria for conducting confirmation tests before embarking on data analysis.

## Introduction

The infectious acute respiratory disease named COVID-19 ^1^ has raised international concern during 2020. It is caused by the novel severe acute respiratory syndrome coronavirus 2 (SARS-CoV-2) ^2^. The first outbreak was confirmed in China in December 2019 ^3^. This outbreak led to 4,642 deaths among 84,149 overall confirmed cases in China, as of April 9, 2020 ^4^. SARS-CoV-2 spread exponentially in China and throughout the world, and the World Health Organization officially declared COVID-19 as a pandemic on March 11, 2020 ^5^. Over 80 million cases were confirmed across the world on December 29, 2020, and about 1.75 million deaths were reported in the same day worldwide ^5^. Countries around the world have worked to prevent the spread of this novel infectious disease. COVID-19 data have been the basis for several policies. Theoretically, data analysis aims to state actual circumstances or reveal new perspectives. However, it is difficult to achieve this objective with COVID-19^6^. A previous study pointed out that differences in testing could mask the actual circumstances of this novel infectious disease ^7^. One way to solve this problem is to use a mathematical modelling of SARS-CoV-2 infection, especially by using the basic reproduction number to survey COVID-19 ^8,9^. Primary statistical data analysis is still the basis of our knowledge on COVID-19, as well as the basis of establishing mathematical models on this disease. Our preliminary research revealed that Japan already had an advanced surveillance system before the pandemic, implemented in line with the Japanese domestic law known as the “infectious disease control law” enforced since 1998 ^10^. In brief, when a patient is suspected to have an infectious disease, such as COVID-19, the doctor must report this case immediately to the Public Health Center (PHC), which then shares the information with the corresponding institutions and the Ministry of Health, Labour and Welfare (MHLW) ^10, 11^. If the suspected patient is confirmed to have an infectious disease, he/she is immediately isolated to prevent spreading the infection, and PHC then identifies the people who were in close contact with the patient, treats them similarly to those who were exposed with the infectious pathogen, and further monitors them to prevent spreading the disease. All data were collected systematically from the National Epidemiological Surveillance of Infectious Disease (NESID) system, as per the law. For example, data on hospitalized COVID-19 patients who underwent additional PCR test to confirm recovery for discharge were not included ^10,11^. In Japan, the first few SARS-CoV-2-positive confirmed cases were reported on January 16, 2020 ^11,12^, with the number of cases increasing almost daily. The Japanese government declared a state of emergency in seven prefectures (Saitama, Chiba, Tokyo, Kanagawa, Osaka, Hyogo, and Fukuoka) on April 7 2020, then this declaration was expanded to the remaining 40 prefectures in April 16, 2020 ^13^. This declaration was gradually lifted by May 25, 2020 ^14^.

This study aimed to investigate the actual situation in the wake of SARS-CoV-2 infection in Japan during 2020. Official data released daily by the MHLW in Japan since January 22, 2020 were analyzed. We found that this data collection system gradually would not work starting at the middle of June 2020. This study pointed out the risks in changing the criteria about data collecting first. Then we conducted machine learning (ML) to estimate the SARS-CoV2 infection rate of Japan and the number of those exposed to the novel virus.

## Methods

### Data analysis

Data were obtained from official announcements of the MHLW in Japan ^11^ and for the seven prefectures ^14-20^. Because MHLW official data represent the Japanese government official announcements about COVID-19, we simply refer to these data as “government data” henceforth. Cases confirmed using PCR tests or SARS-CoV2 antigen tests (simply referred to as “tests” in this study) and SARS-CoV-2-positive patients (simply referred to as “patients” henceforth) were reported from January 22, 2020 ^11^. Therefore, we recorded the overall Japanese data from January 22 until December 31, 2020. The government data were collected by the NESID system, thus the test of government data will be called as tests in law. In contrast, the test data of the seven prefectures were counted in each prefecture (we call them as tests in all-cases). Therefore, these two data are basically different.

### Preliminary steps

Positive rate in law was defined as (number of patients) / (number of tests in law), whereas positive rate all-cases was defined as (number of patients) / (number of tests in all-cases) As all data were recorded in Japanese; we prepared a data archive in English, which is included in the supplemental file. R language was used for data analysis and graph preparation. Sample codes for graphs are listed in the supplemental document.

### Machine learning for estimating the positive rate of SARS-CoV2 infection

Machine learning (ML) was conducted to estimate both the number of tests in law and the positive rate in law in Japan, or each prefecture, during 2020. Briefly, we obtained the regression function by ML, which estimated the number of tests in law from the data of tests in all-cases. We built multi-layer neural networks to work out a supervised learning using R language. We have described ML more precisely in the supplemental methods. Once we obtained the results of estimating the number of tests in law (called tests estimated), the positive rate was estimated, defined as (number of positive confirmed) / (number of tests estimated). Then, we calculated the number of people who might have been exposed to SARS-CoV-2 (referred to as exposed) as (number of confirmed positive tests) / (estimated positive rate)

## Results

### Data quality check

Before the data analysis, we checked the data quality. According to the footnote of the official government announcements in late March ^11^, Chiba, Kanagawa, and Osaka Public Health Centers (PHC) had counted many duplicate samples until March 23, 2020. Thus, the date of these three prefectures were excluded one at a time from the official data. Due to this mistake, we had collected all data from Mach 24, 2020 as the first day of the data analysis, except for the analysis regarding the number of tests, patients, and deaths. Government data for May 4, 2020, were never officially reported; thus, the dataset has missing data. These data were treated as blank.

### Primary data analysis

Both the number of tests and test-positive patients increased exponentially from January 15, 2020, the date when the first patients were confirmed test-positive (Figs. 1a, b). Figure 1c shows the positive rate in Japan in 2020. We could observe that there were rapid fluctuations, with the peaks coinciding with Saturdays and holidays (Fig. 1c, Fig. S1). Careful analysis showed that there were fewer tests on weekends and holidays than on weekdays, while the number of patients increased irrespective of that factor. We concluded that such rapid fluctuations could be ignored because they occurred naturally in the data. We could observe the peaks of positive rate in law three times, the first one was during April 2020, the next was from the middle of July 2020, and the last one was from November 1, 2020 (Figs.1a, c). We call these three peaks the “first wave”, “second wave”, and “third wave,” respectively (Figs. 1a, c). The peak of the first wave was approximately 13%, the second wave was over 5%, and the third wave could not be determined because the ratio was increasing from November 1, 2020 until the last day of 2020. We also recognized the rapid decrease of positive rate in law on June 18, 2020, the positive rate in law has been similar to the positive rate of all cases since the rapid decrease (Fig 1c).

**Fig. 1.**
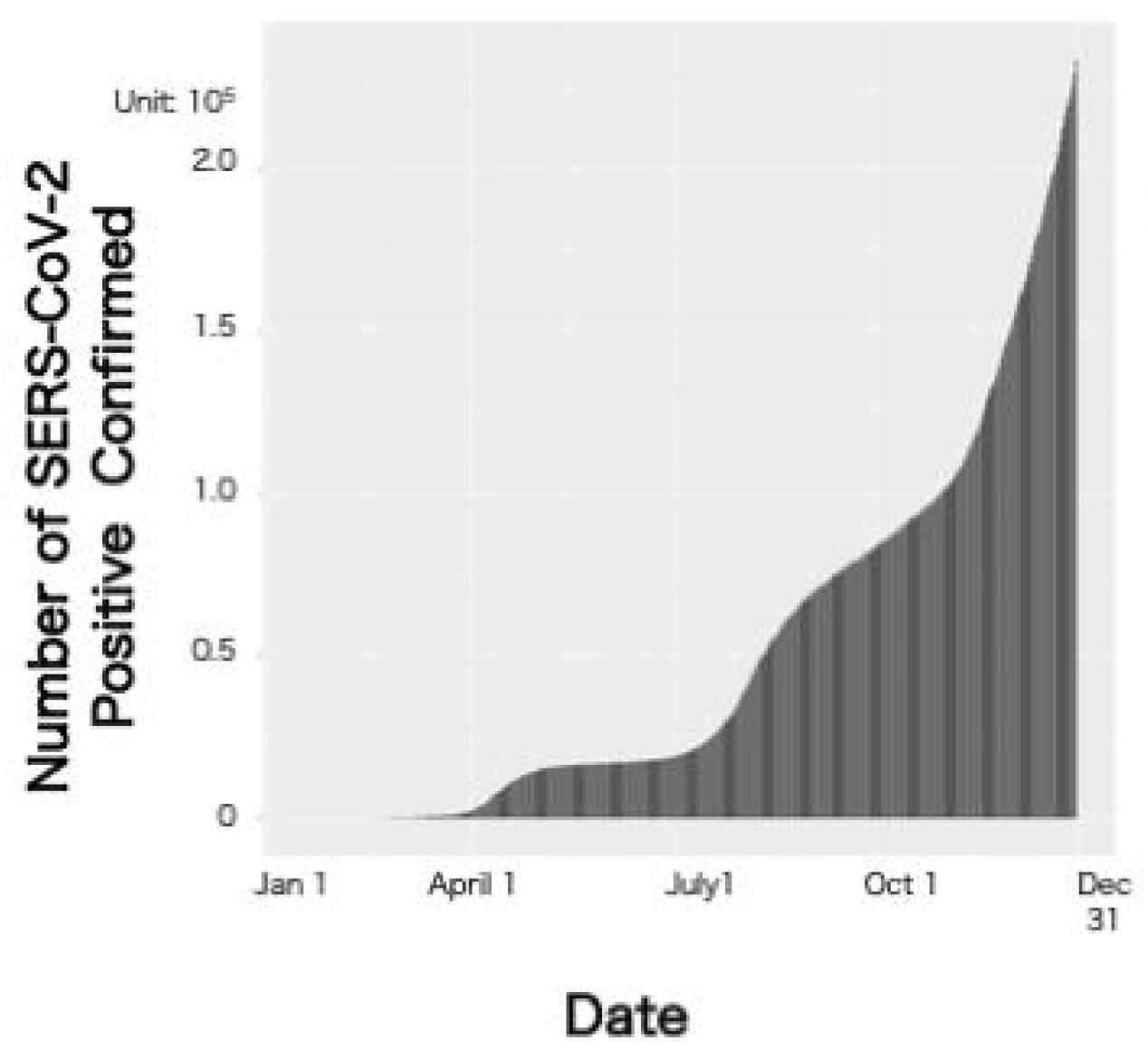
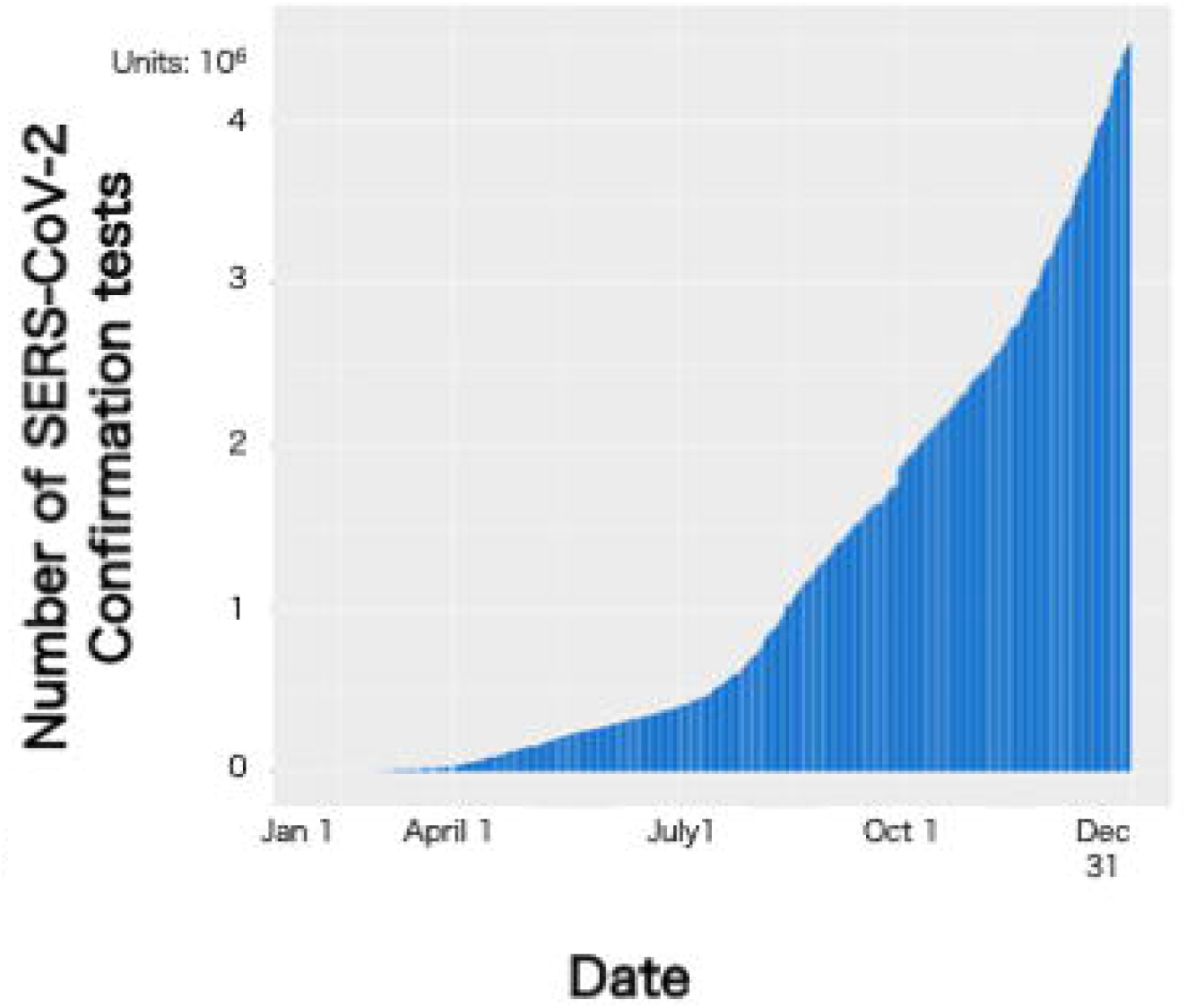
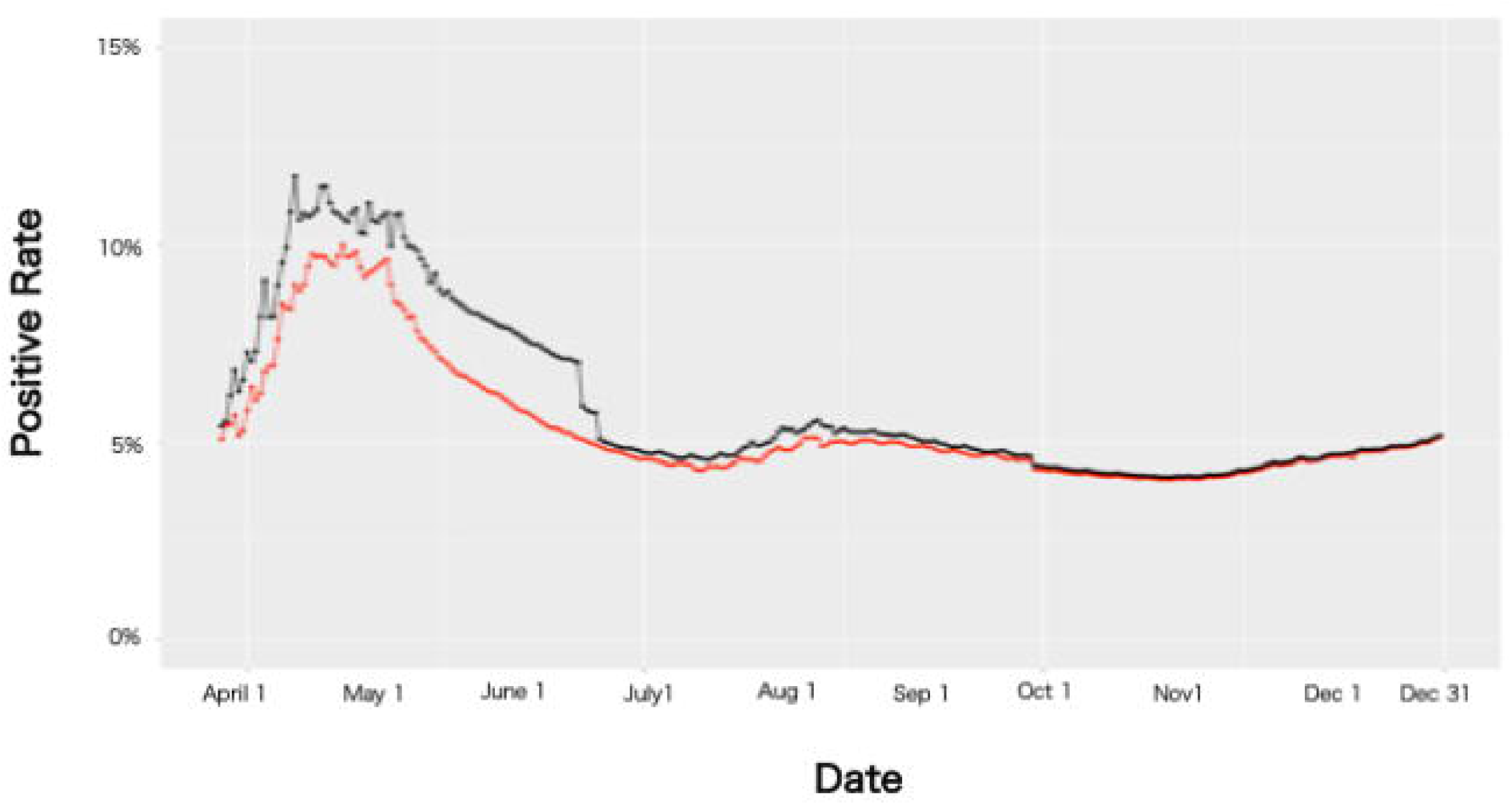
Graphs of COVID-19 confirmation tests (Tests), COVID-19-positive cases (Patients), and Positive Rate in Japan during 2020. (a) Number of patients since the day the first SARS-CoV-2-positive case was confirmed; this number has been growing exponentially. (b) Number of tests since the day the first SARS-CoV-2-positive case was confirmed. The number has been growing exponentially as well. (c) Positive rate in Japan from March 23, 2020 until December 31, 2020. There were many rapid increases and decreases (fluctuations) in a short period. All fluctuations have no sense in this study. Sudden decrease was observed on June 18, 2020.

We further investigated why the rapid decrease occurred. The government official data announced that data collection criteria was changed on June 17, 2020 ^11^, and the data for Tokyo were collected via the NESID system before that day. After that time, the definition of tests included all cases where COVID-19 was confirmed by testing. Due to the change in criteria, the number of tests increased from 16,352 to 61,112 per “single day” in Tokyo. Such a rapid increment was not observed elsewhere in this study in a single prefecture per day ^11,15^. This led to the drastic increase over Tokyo as well as Japan, and positive rate in law rapidly decreased and approached the positive rate all-cases (Figs.1c, 2a). According to the government official announcement on June 18, 2020 ^11^, the data of 19 prefectures, including Tokyo and Osaka, were not guaranteed to be collected through NISID system and were overestimated from the actual number of tests in law. Before that time, because both the number of tests and patients in Tokyo and Osaka were the highest in Japan ^11^, the changing criteria of these two prefectures (or maybe 19 prefectures) made the positive rate in law close to the one using tests for all cases; thus, these results led to weakening of the data analysis in terms of reflecting the real time situation of the novel virus infection.

### Estimating the SARS-CoV2 infection in Japan by using machine learning

As previously mentioned, the government data from June 18, 2020 were less reliable than the ones before that time. One way to solve this problem was to estimate the number of tests and positive rate via a machine learning (ML) method. ML has many parameters that affect the results. Parameter tuning and choosing the proper training data are essential in ML ^21^.

Before ML, we checked the data quality in the seven prefectures. This quality check revealed that Kanagawa datasets were not suitable for daily surveillance of patients and tests (Fig. S2). Therefore, we decided not to use the Kanagawa datasets in this study. This check also revealed that the data of Hyogo data was almost the same as the government official announcement data ^11, 17^. Thus, there was no need to conduct ML for Hyogo data. We found that the Saitama data were always collected under the NESID system ^11^. Thus, we decided to use the government data through 2020 in the two prefectures. According to a footnote on government data ^11^, Chiba and Osaka data are less reliable since April 29, 2020, as well as the data for Tokyo and Fukuoka since June 18, 2020. Therefore, we used the data collected before that day as training data for ML. We first conducted the ML to estimate the number of tests (simply referred to as “estimating” henceforth) of Osaka and Tokyo data, then applied it to the rest of two prefectures (Chiba and Fukuoka), and finally Japan overall.

We first calculated the estimated positive rate for Tokyo. As previously mentioned, we saw that the Tokyo positive rate in law suddenly dropped on June 18, 2020. We obtained the estimated Tokyo positive rate during 2020 from ML, shown in Fig. 2a. We found that there were three peaks during 2020, whose periods were almost the same as the Japan positive rate in law (Fig. 1c).

**Fig. 2.**
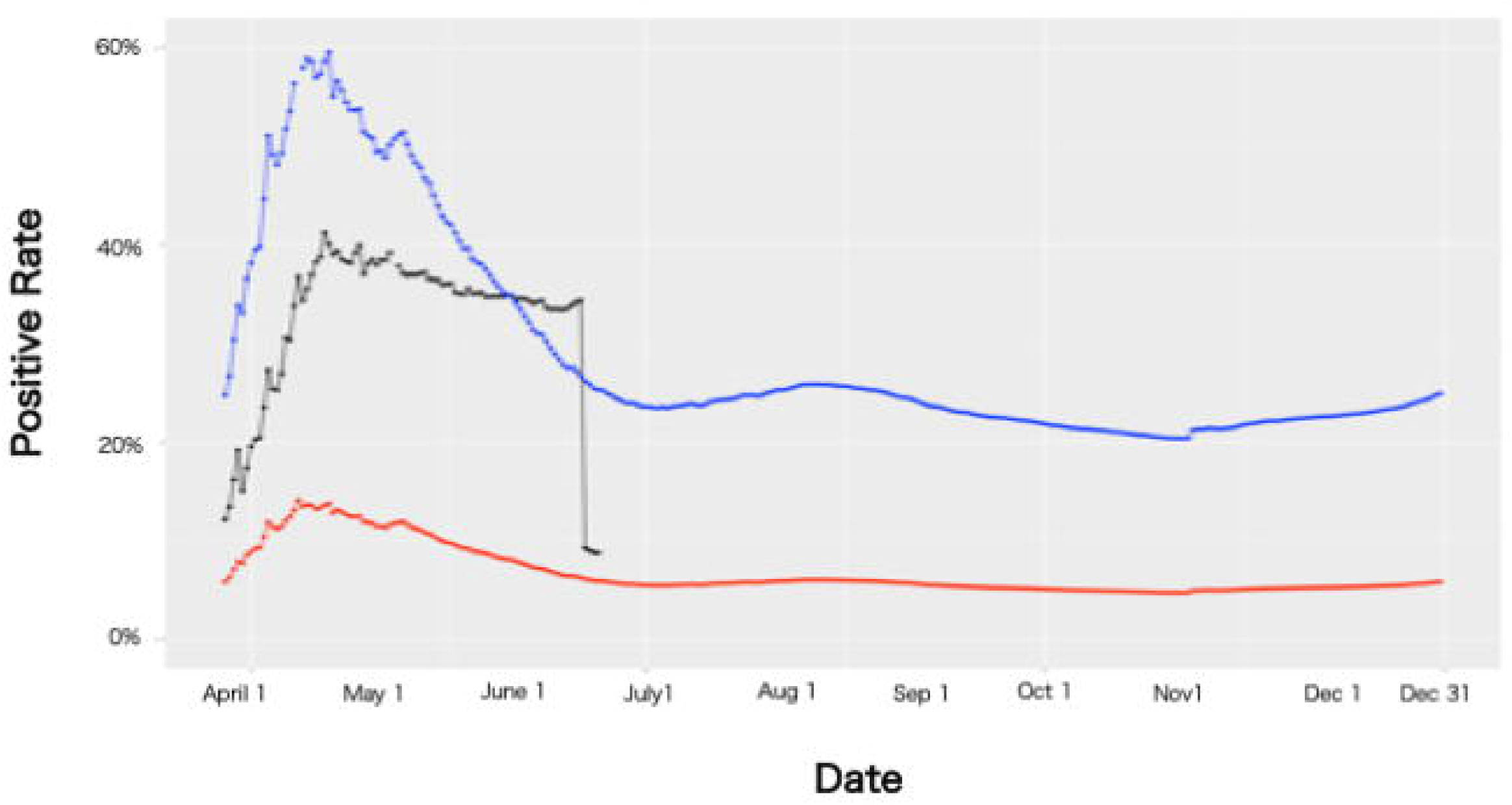
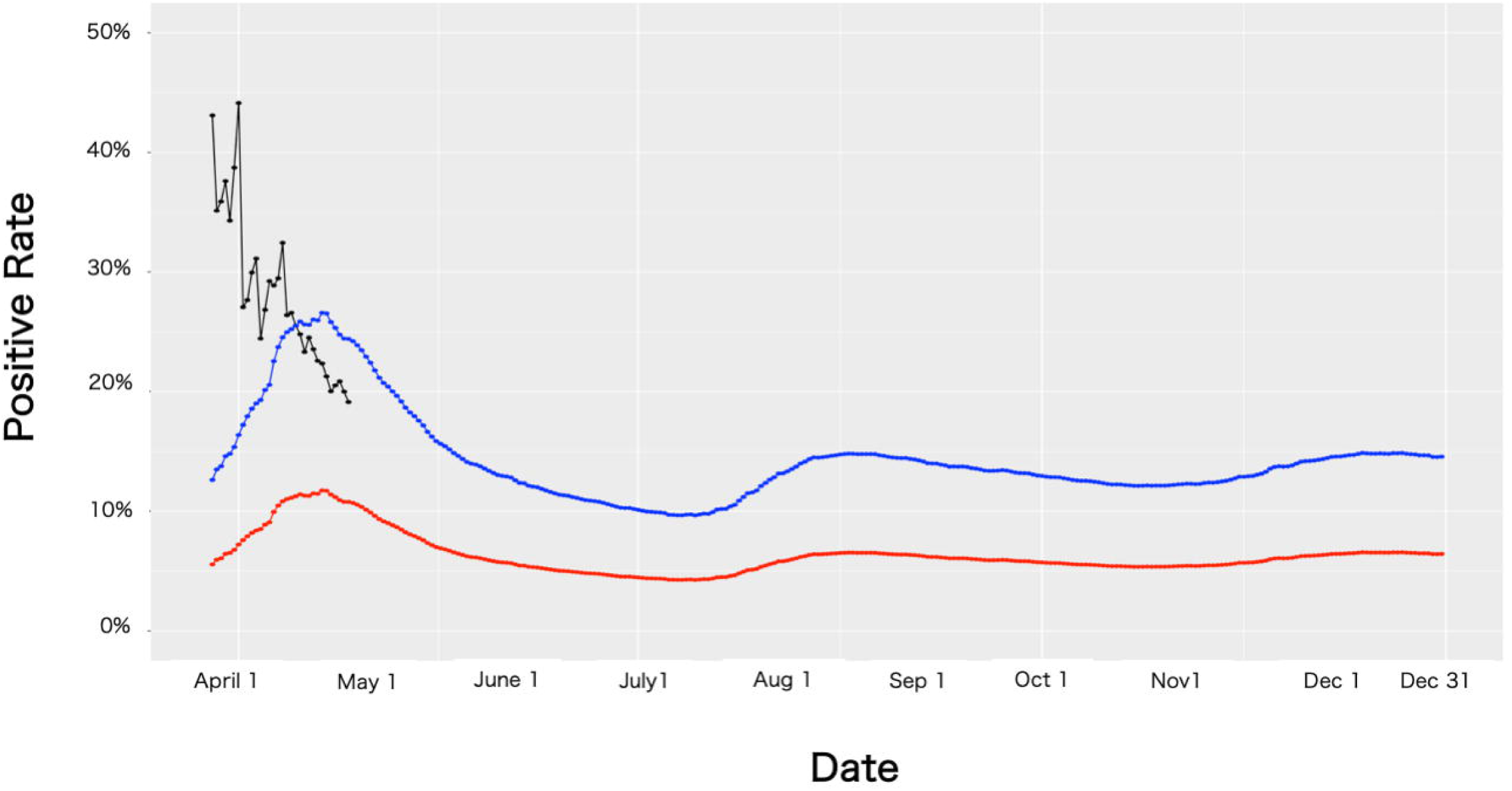
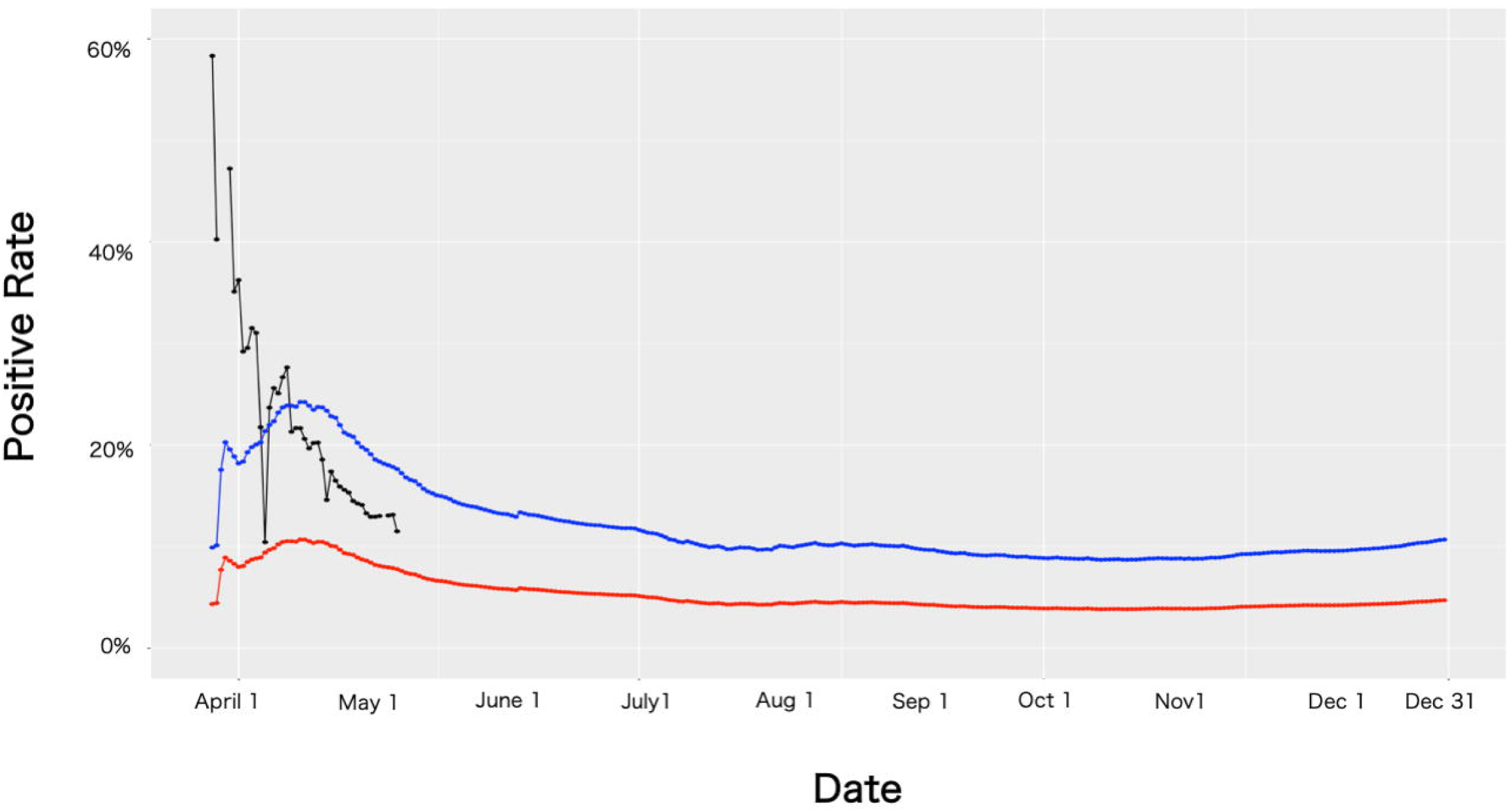
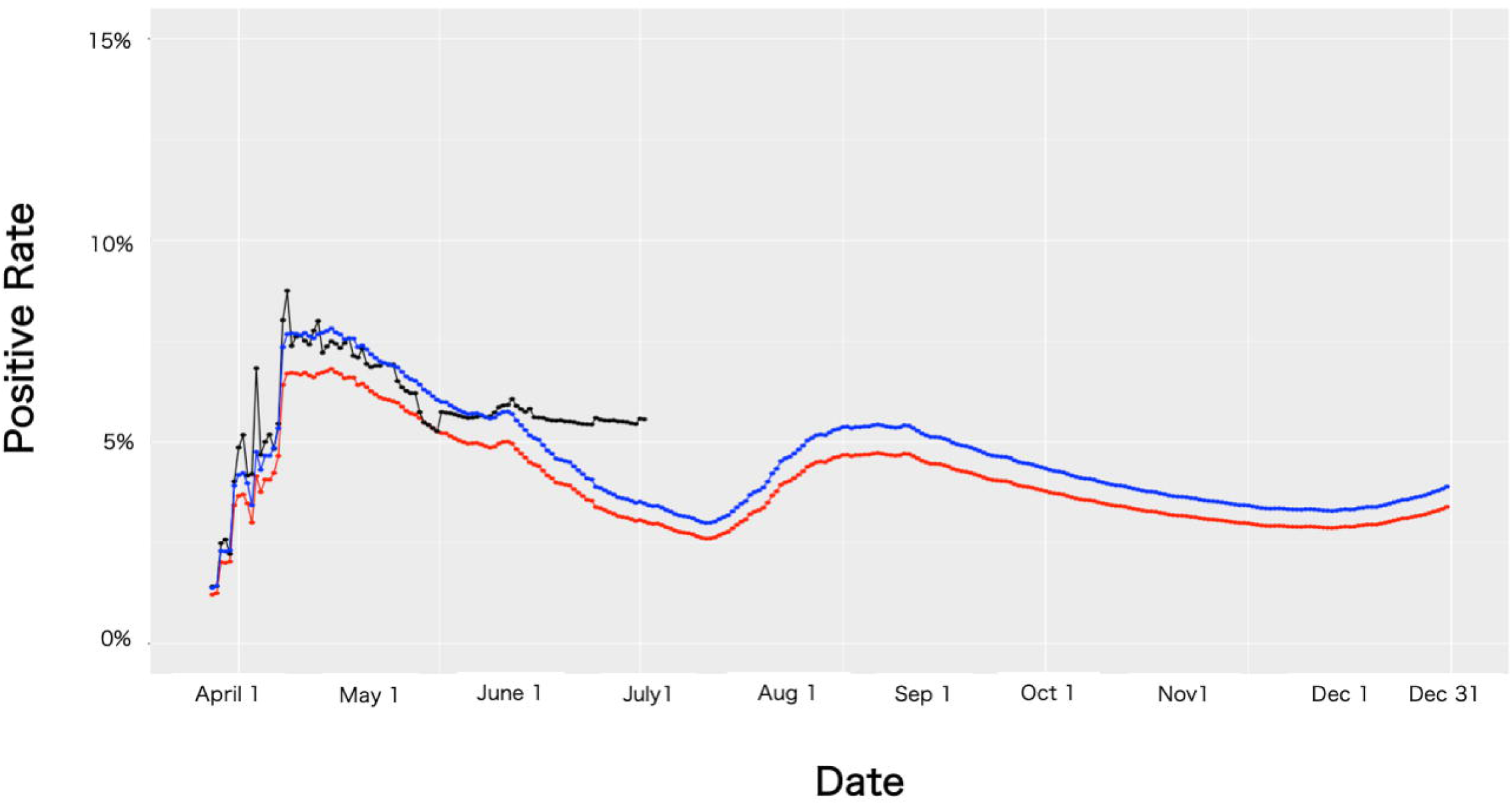
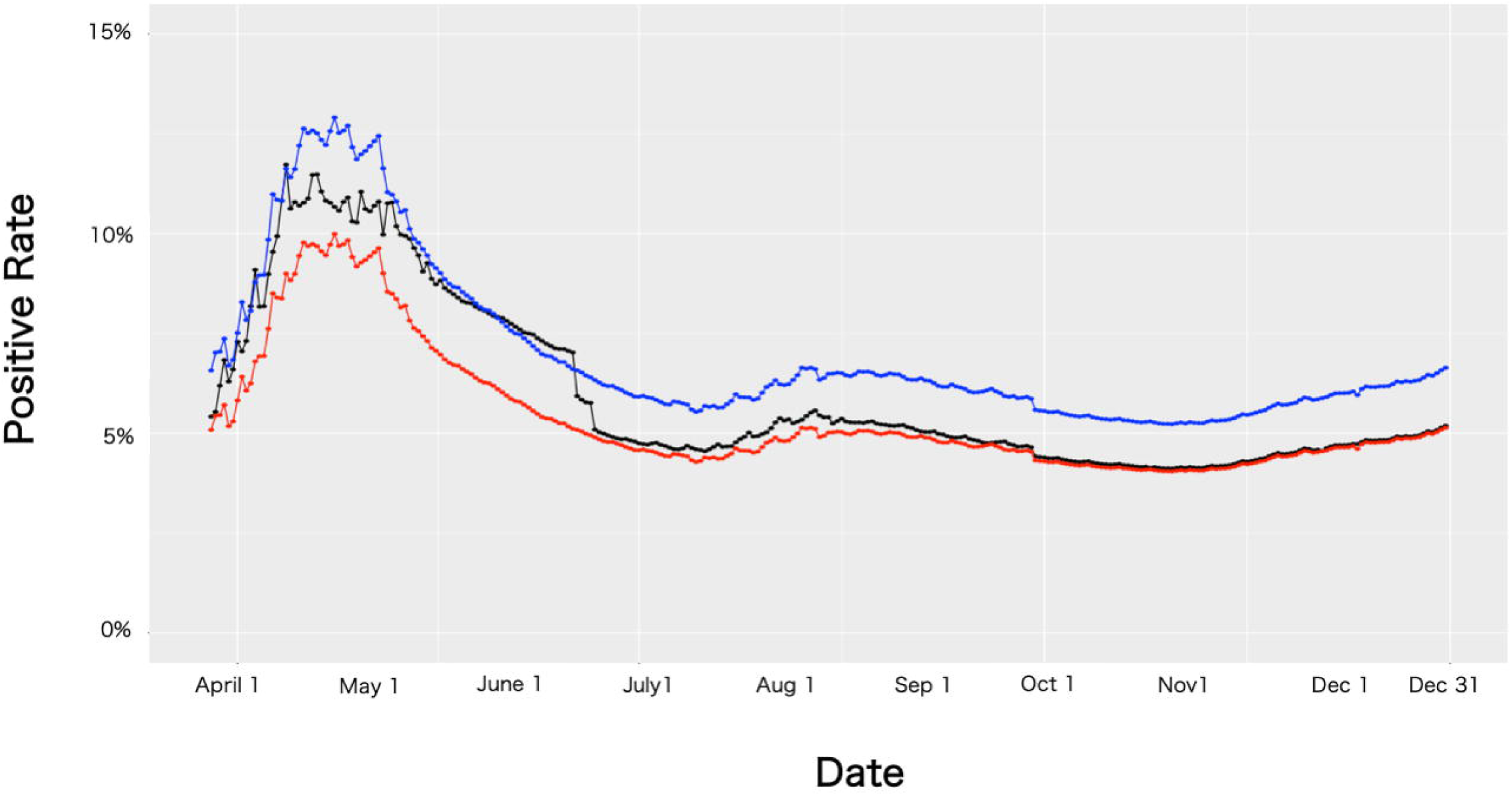
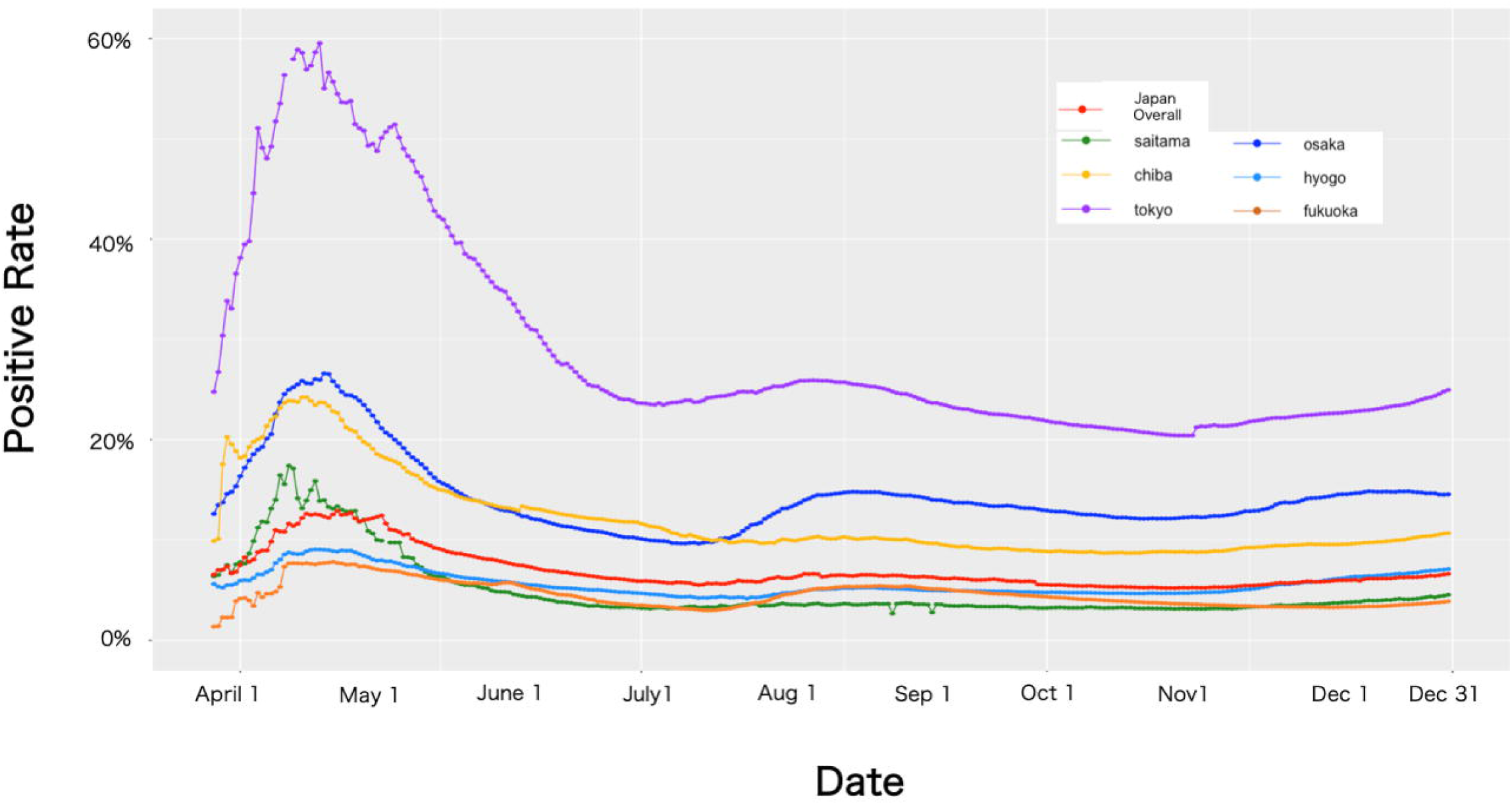
Comparison among the positive rate obtained by the data collected by law, the data counting all confirmation test conducted, and data estimated by machine learning (ML). We compared the positive rate obtained by using the data collected by law (positive rate in law; black lines), the data counting all confirmation test conducted (positive rate in all cases; red lines), and the estimated positive rate obtained by ML (blue lines). (a) Comparison of Tokyo positive rate. The positive rate in law (black line) drops on June 18, 2020 and is almost the same as the one in all cases (red line) since June 21, 2020. The estimating positive rate was always over 20% during 2020. (b) Comparison of Osaka positive rate. The tests were counted in duplicate until March 23, 2020, at Osaka; thus, the positive rate in law is not stable in this survey. We observed three peaks during 2020. (c) Comparison of Chiba positive rate. The tests were counted in duplicate until March 23, 2020 at Chiba and Osaka, thus, positive rate in law is not stable. We observed two peaks during 2020 in contrast to Osaka data. (d) Comparison of Fukuoka positive rate. While positive rate in law stayed stable during June 2020, the estimated positive rate decreased as days passed through the same period. (e) Comparison of Japan overall positive rate. The positive rate in law (black line) dropped on June 18, 2020 and was almost the same as the one in all-cases (red line) since June 21, 2020 as well as in Tokyo. In contrast, the estimating positive rate seemed to be stable or to decrease gradually in this period. (f) Comparison of Japan overall, Tokyo, Osaka, Chiba, and Fukuoka estimated positive rate, and Saitama and Hyogo positive rate in law. All seven graphs indicate higher percentage until May 2020 than the rest in 2020. Rates were increasing from November 2020 in seven graphs. These seven graphs indicate the same tendencies except for the period between the middle of August and September.

Through parameter tuning, the error of number was found to be within 1% (a part of data is shown in the supplemental document). The first wave seemed to terminate at the end of May 2020, then the rate converged to approximately 23%-30%. The second wave also seemed to settle down in the middle of September 2020, then the rate converged from 20% to 25%. In conclusion, SARS-CoV-2 infection in Tokyo was estimated to converge from 20% to below 25% after the first wave of spread of the virus, and there were three major periods wherein the novel virus spread. Figures 2b, c, d shows the estimated positive rate in Osaka, Chiba, and Fukuoka, respectively. A similar trend can be observed, except in Chiba, where the second wave was not observed. Figure 2e shows the comparison between the three positive rates in Japan. Estimated positive rate in Japan vanished during the rapid decrease on June 18, 2020, and then stayed higher than the other two positive rate. There were three infection-spreading periods observed, which is coincident with the data of each prefecture. The first wave seemed to terminate at the end of May 2020, and the rate converged over 6% to around 8%. The second wave settled down around the middle of September 2020, and the rate was converged in around 7%. The third wave seemed to start on November 1, 2020 until the end of the investigation period. Figure 2f shows the comparison of Japan, Tokyo, Osaka, Chiba, and Fukuoka estimated positive rate, and Saitama and Hyogo positive rate in law. The data showed that there were at least two peaks during 2020, the first peak was from the first day of the observation until almost May 31, 2020, and the last one started on November 1, 2020. The rate was converged between June 1, 2020, until the middle of August, and between the middle of September and October 30, 2020.

### Estimation of the overall number of people who may have COVID-19 and deaths related to COVID-19 during 2020

As previously discussed, we observed that there were peaks in SARS-CoV-2 infection, and the estimated positive rate between the first and second wave converged to approximately 6%-8%; the one between the second and third wave converged to around 7% (Fig. 2f). Therefore, Japan’s SARS-CoV-2 infection rate before the third wave was estimated to be between 6% and 8%. Based on this result, and the data on Japan’s confirmed positive results, we computed the number of individuals exposed, shown in Fig 3a. The number of exposed individuals grew as days passed. On June 16, 2020, the number of exposed individuals was about 0.27 million; on September 20, 2020, it was 1.37 million; and on October 31, 2020, it was 1.95 million.

**Fig. 3.**
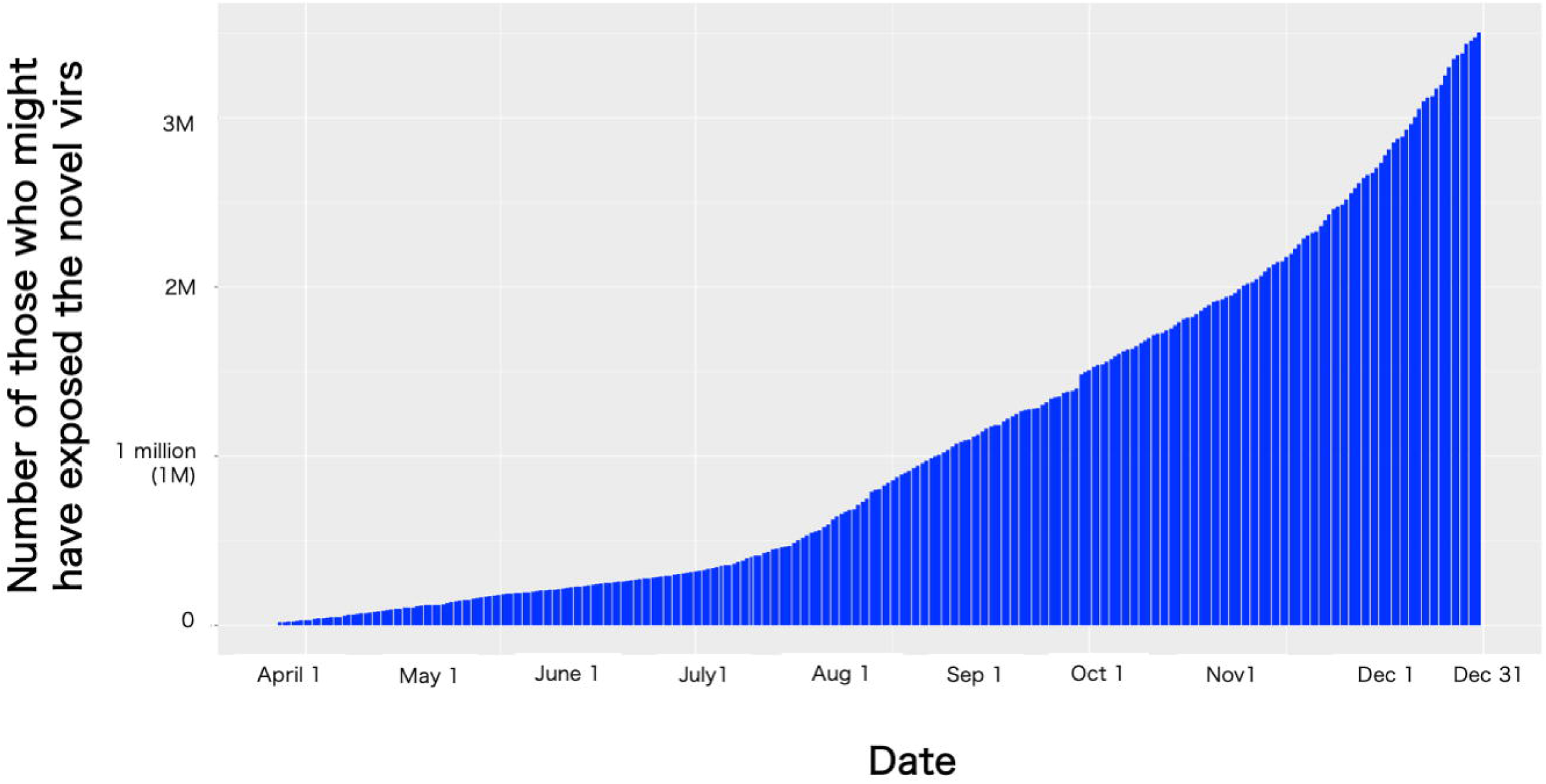
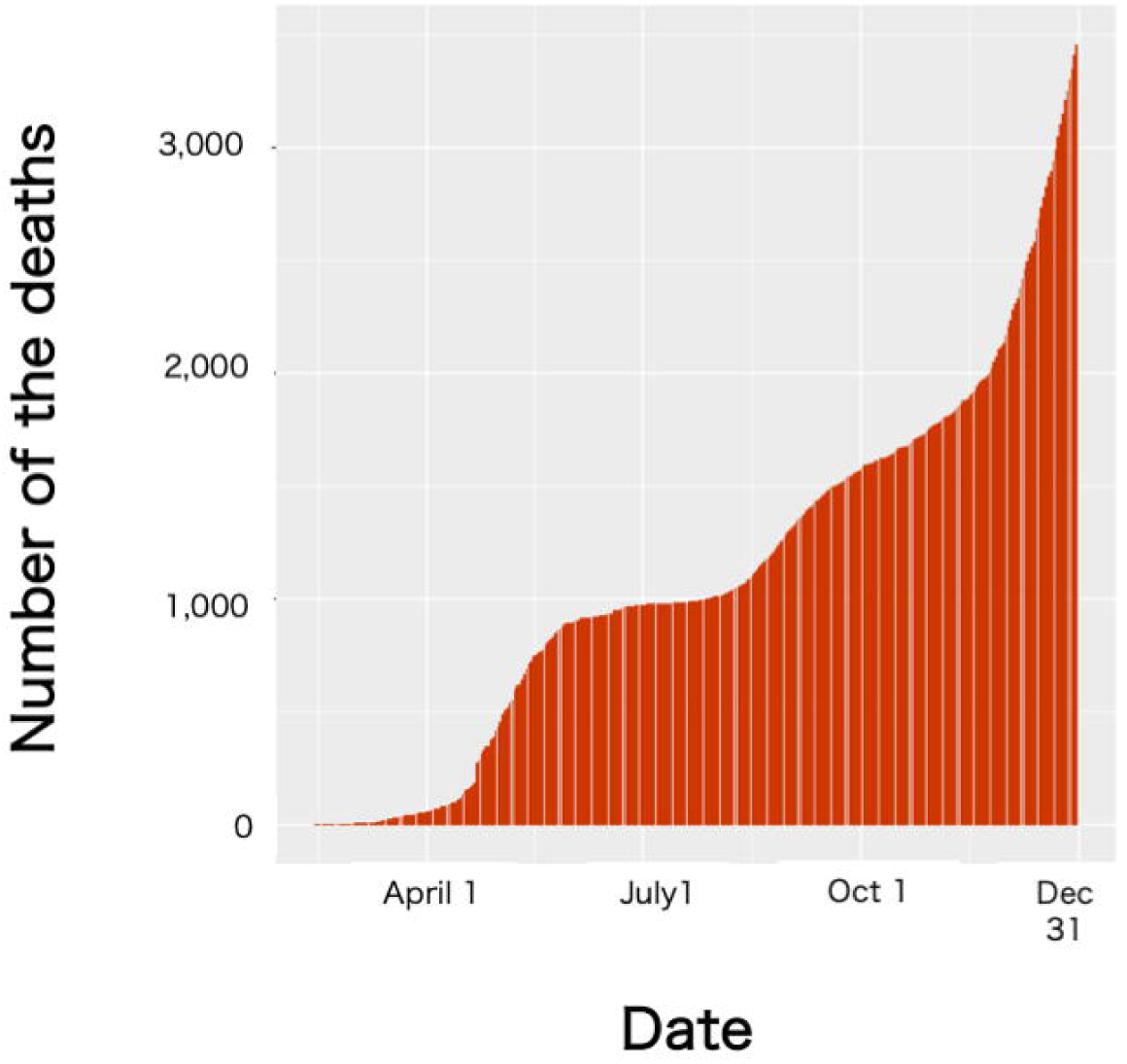
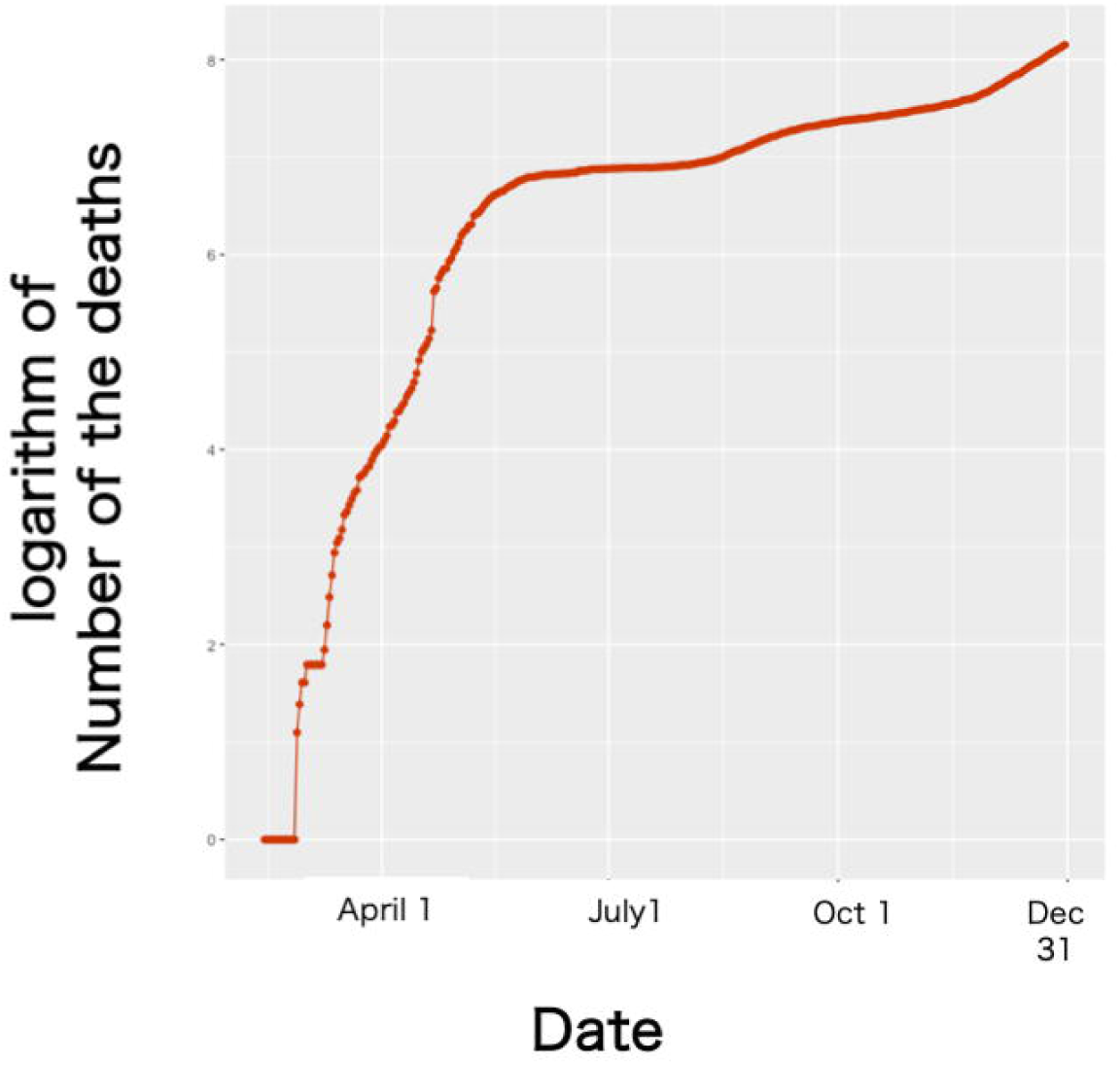
Estimating number of those who might have been exposed to the SARS-CoV-2 and COVID-19-related deaths. (a) Estimated number of those who might have been exposed to the SARS-CoV-2. (b) COVID-19-related deaths since the day the first COVID-19 death was confirmed. (c) Logarithm of the number of deaths. The Y-axis shows a logarithmic scale. The slope is almost zero from the June 2020 until October 2020, which suggests that the growth of deaths during this period is not exponential, but linear.

Next, we focused on the deaths related to COVID-19 in Japan during 2020. The first death related to COVID-19 was reported on February 14, 2020; then, the deaths increased almost daily, and reached 1,000 on July 28, 2020, at over 2000 on November 24, 2020, and over 3,000 on December 23, 2020 (Fig. 3b). If we assumed that the number of deaths increased exponentially as the day passed, then the slope of the logarithm of number of deaths determines its characteristics. Figure 3c shows the logarithm of number of deaths. We found two rapid slopes; the first one was until around the end of May, and the second rapid slope was from November 1, 2020, suggesting that the infection of the novel coronavirus occurred during these periods in Japan. The logarithms from June 2020 until October 31, 2020 were calculated as almost the same (Fig. 3c), suggesting that the increment in deaths during this period was not exponential but linear.

## Discussion

We observed that the NESID system facilitated data analysis regarding COVID-19 in Japan with some problems. This system was designed to be able to trace suspected people who might have been exposed to an infectious pathogen like SARS-CoV-2 and identify the spreader, i.e., some persons in an area or areas wherein infection was spreading. This system provides us the estimated positive rate and the number of exposed individuals. It was regrettable, however, that the definition for counting cases changed in Osaka from May 2020 as well as in the other 18 prefectures, including Tokyo, from June 17, 2020.As both the numbers of tests and patients in Tokyo and Osaka were the highest in Japan ^11^, the exclusion of these two prefectures would have weakened data accuracy according to the NESID system. The reason why such a change occurred is that PHC do not have enough capacity to engage COVID-19 business with routine business ^22^. The positive rate in law and the all cases differed markedly in Japan as well as in Tokyo (Figs. 1c, 2a, e), and this difference led to contrasting conclusions: data analysis using different definitions can mask real-time situations. A previous study suggested that different methods of data analysis could mask the real events surrounding COVID-19 ^7^. However, the data sample was limited in the former study. In this study, data analysis highlighted the importance of the definition of the number of tests conducted. Therefore, this study suggests that the method of data collection must be precisely defined before data analysis.

To overcome this problem, we conducted ML to estimate the SARS-CoV-2 infection rate in Japan in 2020. We found that Japan had at least two peaks of the novel virus infection. After the first wave seemed to settle down, Japan’s positive rate was estimated to be between 6% and 8%; then, the number of exposed was estimated to be approximately 0.27 million. The positive rate after the second wave was estimated to be around 7%, and around 1.95 million people were estimated to be exposed to the novel coronavirus. Although we can see that Japan was in the third wave in the last two months of 2020, we cannot predict when the peak of the third wave occurs. Thus, we cannot estimate the number of people who might have COVID-19.

The mortality data related to COVID-19 were also analyzed. We found that the deaths increased exponentially until May, 2020 and between November and December 2020, while the deaths increased linearly from June 2020 until October 2020. Based on this analysis, we concluded that there were at least two infection spreading periods in Japan, the first one was from the day that the first COVID-19 patient had been reported until May 2020 and the second one was from November 1, 2020. The infection rate of the novel virus during 2020 was estimated to be around 6%-8% in Japan. Note the “infection rate” was obtained by the conclusion of this study, while the “positive rates” were computed by the definition described in the methodology.

In conclusion, Japan had a well-established surveillance system for infectious diseases before the COVID-19 pandemic. Analysis of the data provided from this system revealed that there were two spreading periods of SARS-CoV-2 infection in Japan during 2020, and infection rate was estimated to be between 6% and 8% during 2020. In addition, it also suggested that the criteria for the definition of virus-positive tests should be clearly defined before conducting data analysis.

## Supporting information

all data used in this study

## Data Availability

All data are obtained by governments open data archive.
Because all data used in this study are written in Japanese, the original data are listed in main text and/or supplemental data

## Acknowledgements

The author thanks all those who supported this study. The author also thanks Editage (www.editage.jp) for providing editing assistance for this paper. This report is dedicated to all victims of COVID-19 worldwide.

## Disclosure statement

The author declares no competing interests

## Funding

The author received no financial support.

## Notes

### Competing Interest Statement

The authors have declared no competing interest.

### Funding Statement

The author receives no financial support.

### Author Declarations

All data are obtained by governments open data archive per admission of each government's open data policy.

