## supplemental documents for "Estimation of infection rate and the population size potentially exposed to SARS-CoV-2 in Japan during 2020"

**Supplemental method**

***Design of machine learning***

To begin with, we decided to use the training datasets for ML through data quality check. The training datasets were separated at random. We then defined ¾ of the datasets as training data for learning, and the rest were defined as test data. We used the R package “caret” for separating the datasets at random.

We constructed the neural network system for supervised learning using R package “keras” ^1^. The detailed code of the ML that we conducted are described next. One of the characteristics of our codes is that it was set sufficiently larger in each layer compared with the input layer’s dimension. This design was intended to minimize training errors ^2^. Training errors are measured by the least square methods.

***The sample code of ML about Japan datasets***

#this is the primary dataset of ML in this code

ML_japan <- Japan_datasets

\# Caret and keras use a random number in their algorithm. The random number can be fixed by this code if you want to reproduce the results of ML.

set.seed(13)

#Creating the training datasets for ML

japan_index <- createDataPartition(y=japan_train_datasets$japan_conducted, p=0.75, list=FALSE )

japan_train <- japan_train_datasets[japan_index,]

japan_test <- japan_train_datasets[-japan_index,]

japanXtrain <- japan_train$japan_overall_conducted

japanYtrain <- japan_train$japan_conducted

japanXtest <- japan_test$japan_overall_conducted

japanYtest <- japan_test$japan_conducted

#Building the neural network by the R package “keras”

japan_model <- keras_model_sequential()

#We set the unit number almost the same as the number of JapanXtrain. If learning fails, changing unit number would lead to success.

japan_model %>%

layer_dense(units=64, activation = "relu", input_shape = 1 ) %>%

layer_dense(units= 256, activation = "relu" ) %>%

#We set the unit number almost the same as the number of JapanXtest.

layer_dense(units=28, activation = "relu") %>%

layer_dense(units=1, activation = "relu")

japan_model %>% compile(optimizer=optimizer_adam(), loss="mse" )

#Output and the plots were generated as shown in Fig. S3.

history_japan <- japan_model %>% fit(japanXtrain, japanYtrain, epoch=50, batch_size=1, verbose=1, validation_data=list(japanXtest, japanYtest ) )

＃Output estimation and collect the data,

est_japan_conducted <- predict(japan_model, ML_japan$japan_overall_conducted )

ML_japan <- tibble(ML_japan, est_japan_conducted)

est_japan_postive_rate <- ML_japan$japan_positive / ML_japan$est_japan_conducted

est_japan_postive_rate <- ML_japan$japan_positive / ML_japan$est_japan_conducted

＃We gained the graph as shown in Figure 2.

ggplot(data=ML_japan, mapping = aes(x=date ) )+

geom_line(mapping = aes(y=japan_positive_rate_all), color="red")+

geom_point(mapping = aes(y=japan_positive_rate_all), color="red")+

geom_line(mapping = aes(y=japan_positive_rate), color="black")+

geom_point(mapping = aes(y=japan_positive_rate), color="black")+

geom_line(mapping = aes(y=est_japan_positive_rate), color="blue")+

geom_point(mapping = aes(y=est_japan_positive_rate), color="blue")+

xlab("Date")+ ylab("Positive Rate")+

＃we change the value of y-axis into the percent style.

scale_y_continuous(labels = scales::percent, limits = c(0,0.15) )

**Supplemental Results**

Theoretically, the supervised learning of regression first computes the regression function from the training data; then, the test data validate the results and modify the parameter of regression function. Repeating this algorithm, we obtain the results of best regression assessing by reducing the training errors. The neural network system, one of the ML methods, is designed to execute this algorithm efficiently ^3,4^. This is the reason why we adapted this method in this study. Figure S3 show the histories of ML, Fig. S3a shows the history of learning success, while Figure S3b shows the failed attempts. If we succeeded in the learning, no matter what the results were, the value of the ML training error dropped as the learning steps increase, and then converged to some value, which was usually unknown (Fig. S3a). If the learning failed, the training error value did not change, right from the first step of the learning (Fig. S3b). We had set the first goal of this study to obtain success in learning, as mentioned above. We further assessed the result of the first goal, which has been mentioned in the main text.

Because training steps are an important parameter of ML ^3,4^, we searched for the optimum number of steps. Figure S3a shows that the training error values were almost stable at 30–50 steps. We conducted training at 100 steps, 1000 steps, and 5000 steps using the Osaka datasets. The differences of the steps are within 1% of the estimating positive rate (data not shown). Keras uses the random number in its algorithm^1,4^. Thus, we conducted ML using the same datasets several times while refreshing the random number. The differences in this trial were also within 1% (data not shown). Therefore, we concluded the range of error in this ML to be 1% of the estimating positive rate.

Finally, we accessed the proper training datasets. We were anxious that the Osaka and Chiba training datasets were too small to obtain good results while designing the ML. Thus, training data for Chiba, Tokyo, Osaka, Fukuoka, and Japan were combined to conduct a new ML for each prefecture and Japan (we called this combined train data as combined train datasets). The results for Tokyo are graphed in Fig S4. We observed that the estimating positive rate for Tokyo were always less than the ones obtained via the NESID system, including during the period between June 18, 2020 and June 20, 2020, where, as we pointed out in the main text, the recording tests criteria was changed. The reason for conducting this ML was that we assumed that the Tokyo data since June 18, 2020 never reflected the actual circumstance of Tokyo; that is, the positive rate must be higher than the obtained results since June 18, 2020. However, the results of ML, using the combined train datasets, conflicts our assumption (Fig.S4). Therefore, we concluded that ML using theses train datasets is not suitable in this study.

**Supplemental figure legend**

**Fig. S1 The positive rate graph highlights Saturdays and holidays.**

The positive rate across Japan during the first half of 2020. Rapid increases and decreases essentially coincide with Saturdays and holidays, which are highlighted with black shadows.

**Fig.S2 Kanagawa positive rate graph.**

The positive rate for Kanagawa is shown. According to the footnote of government official announcement, the Kanagawa Public Health Center counted the sample duplicated until March 23, 2020 and middle of May 2020. This event caused the graph to oscillate widely in the first half of 2020. Although the number of positive confirmed individuals in Kanagawa steadily increased throughout 2020, the number of Kanagawa tests had an unstable transition, i.e., recording numbers were unchanged for about seven days, then they grew rapidly. Because of this event, we found that the positive rate increased gradually for a short time, then decreased greatly within a day. This is because the graph is unusual compared with other prefectures or Japan. Therefore, we decided not to analyze the Kanagawa data in this study.

**Fig. S3 History of Machine Learning (ML)**

Blue lines indicate the value of loss obtained by train data, while the green lines indicate the one obtained by test data. ML was designed to minimize these values.

(a) If ML succeeded, then the value varies as the time passes and is converged.
(b) If ML failed, then the value never changes.

**Fig. S4 Overfitting of Machine Learning (ML)**

Comparison of the Tokyo positive rate, using the data collected by law (shown in black), by counting all-cases (shown in red), and estimating the positive rate (shown in blue). The sky-blue line indicates the result of ML using the combined data from the four prefectures (Tokyo, Osaka, Chiba, and Fukuoka) and Japan overall, while the blue line indicates the result of ML trained by Tokyo data. The sky-blue line is always lower than the black line, which conflicts our assumption.
