## Supplementary figures and images for "Estimation of infection rate and the population size potentially exposed to SARS-CoV-2 in Japan during 2020"

### Supplemental Figure 1

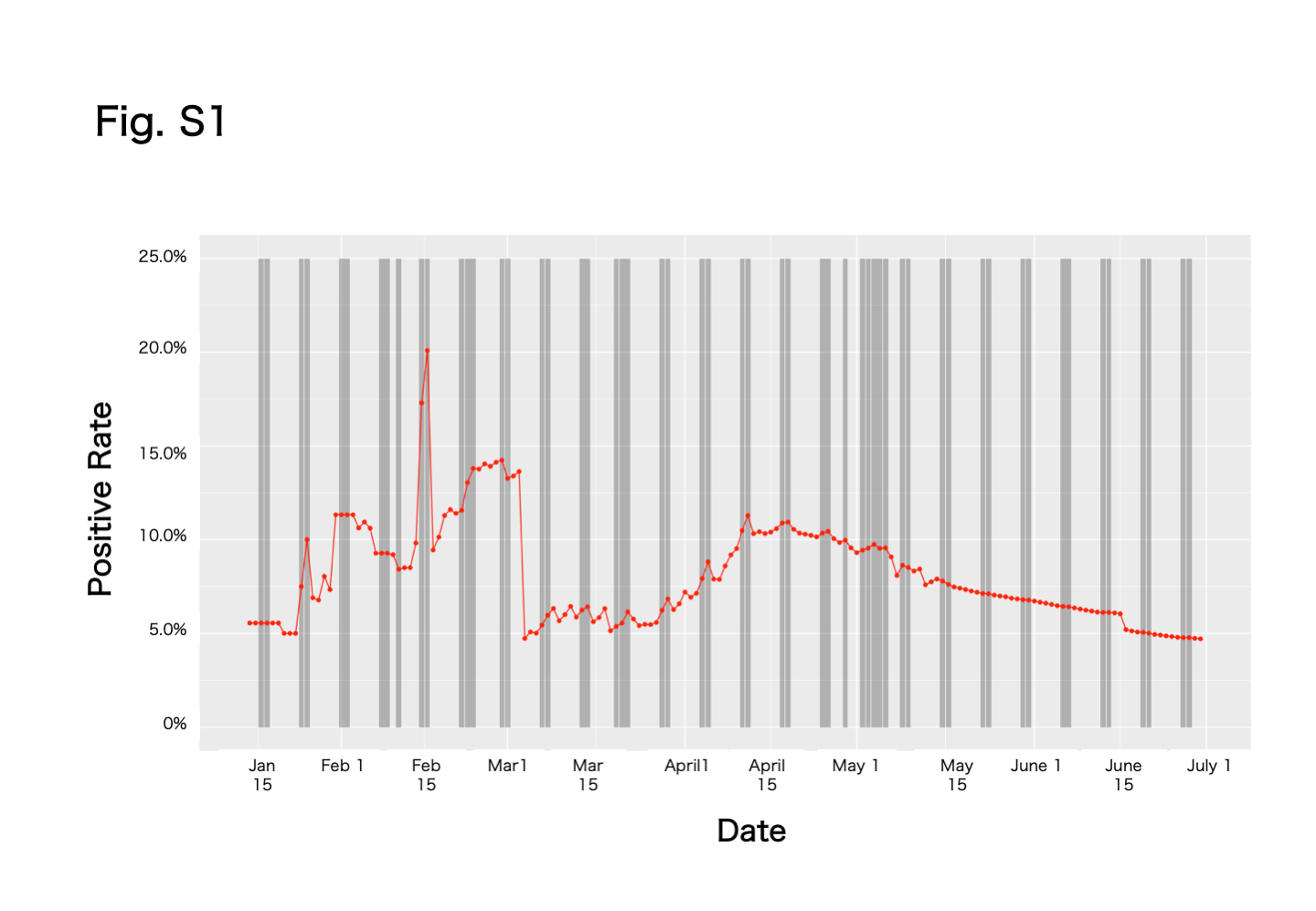

### Supplemental Figure 2

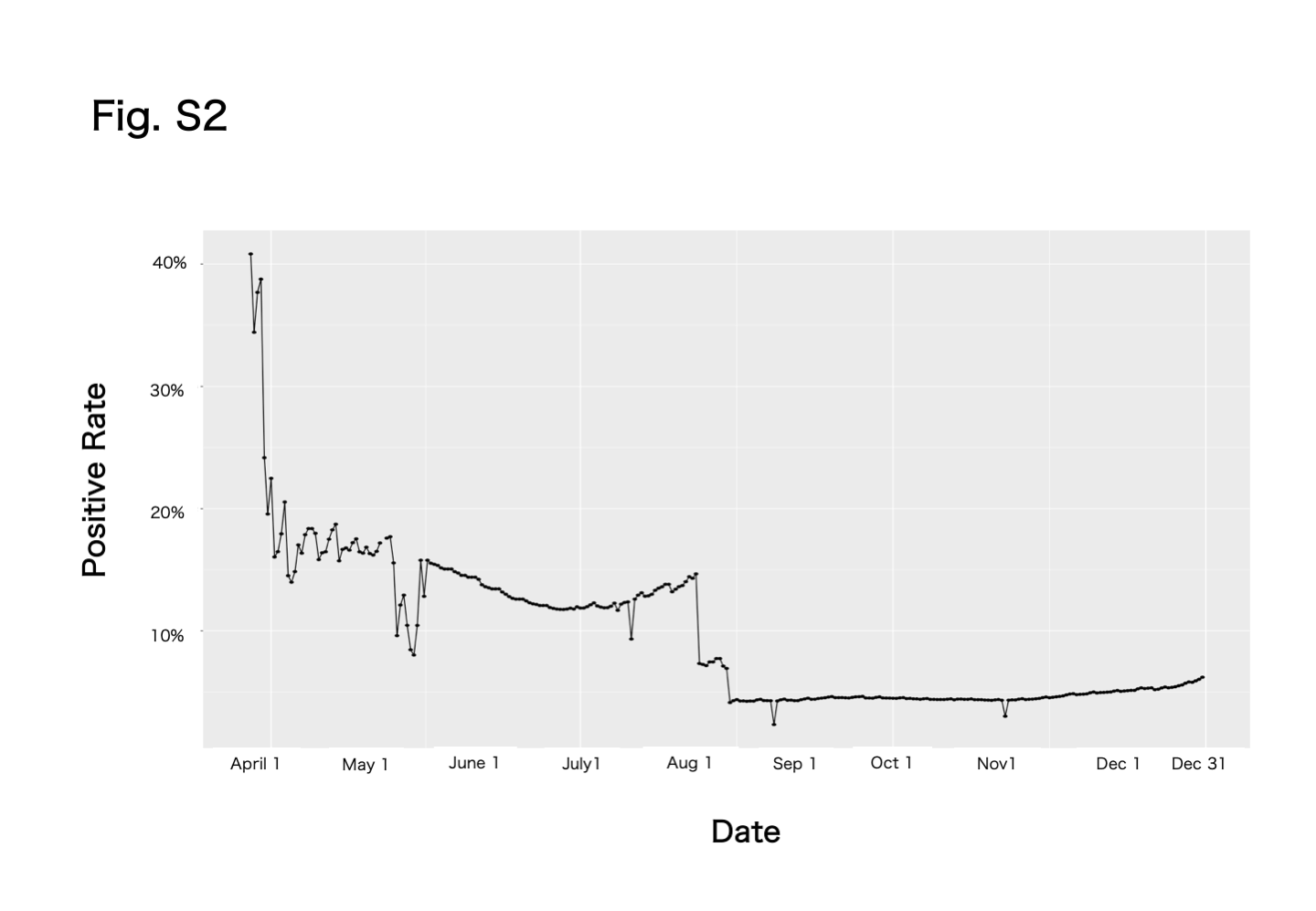

### Supplemental Figure 3

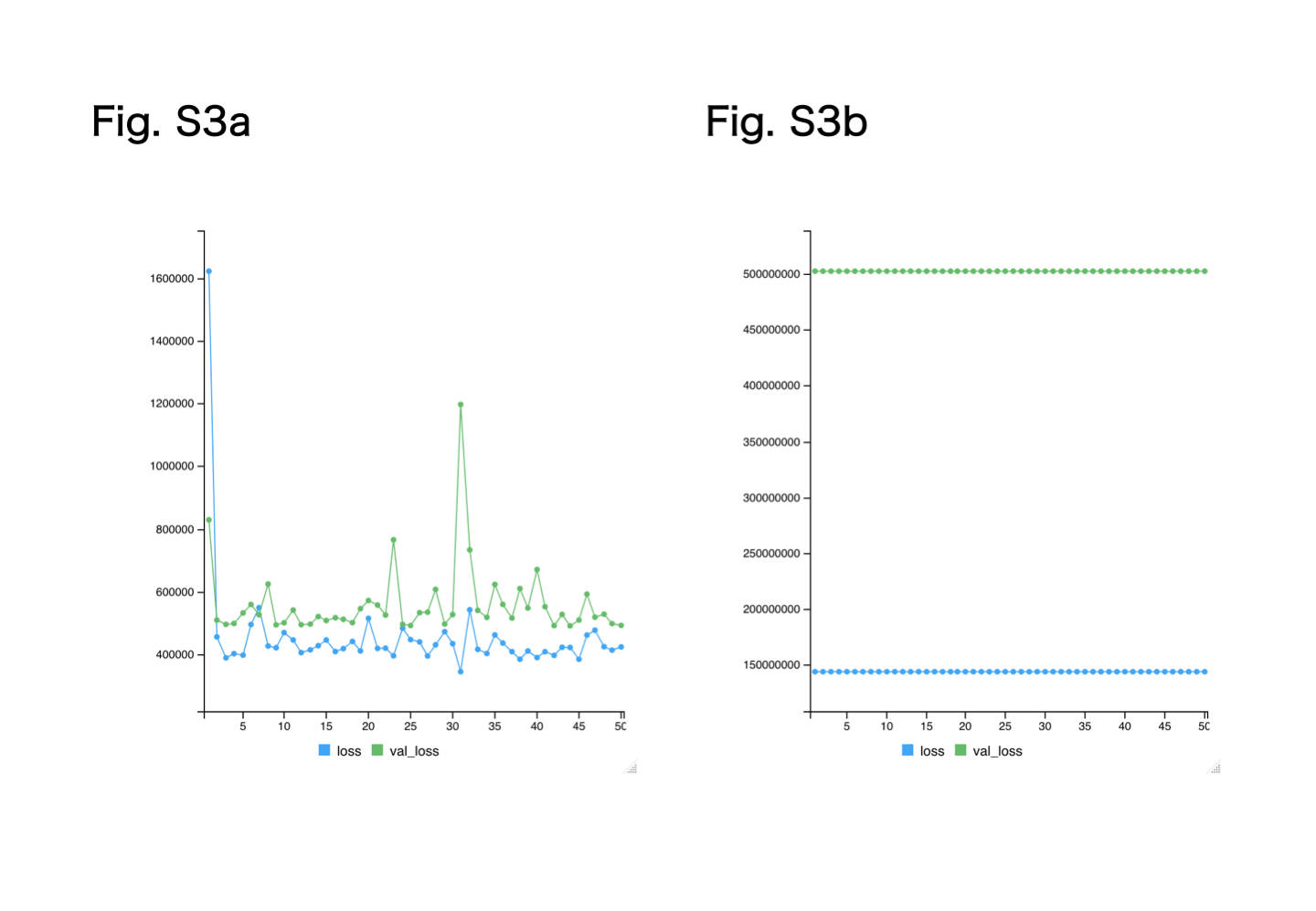

### Supplemental Figure 4

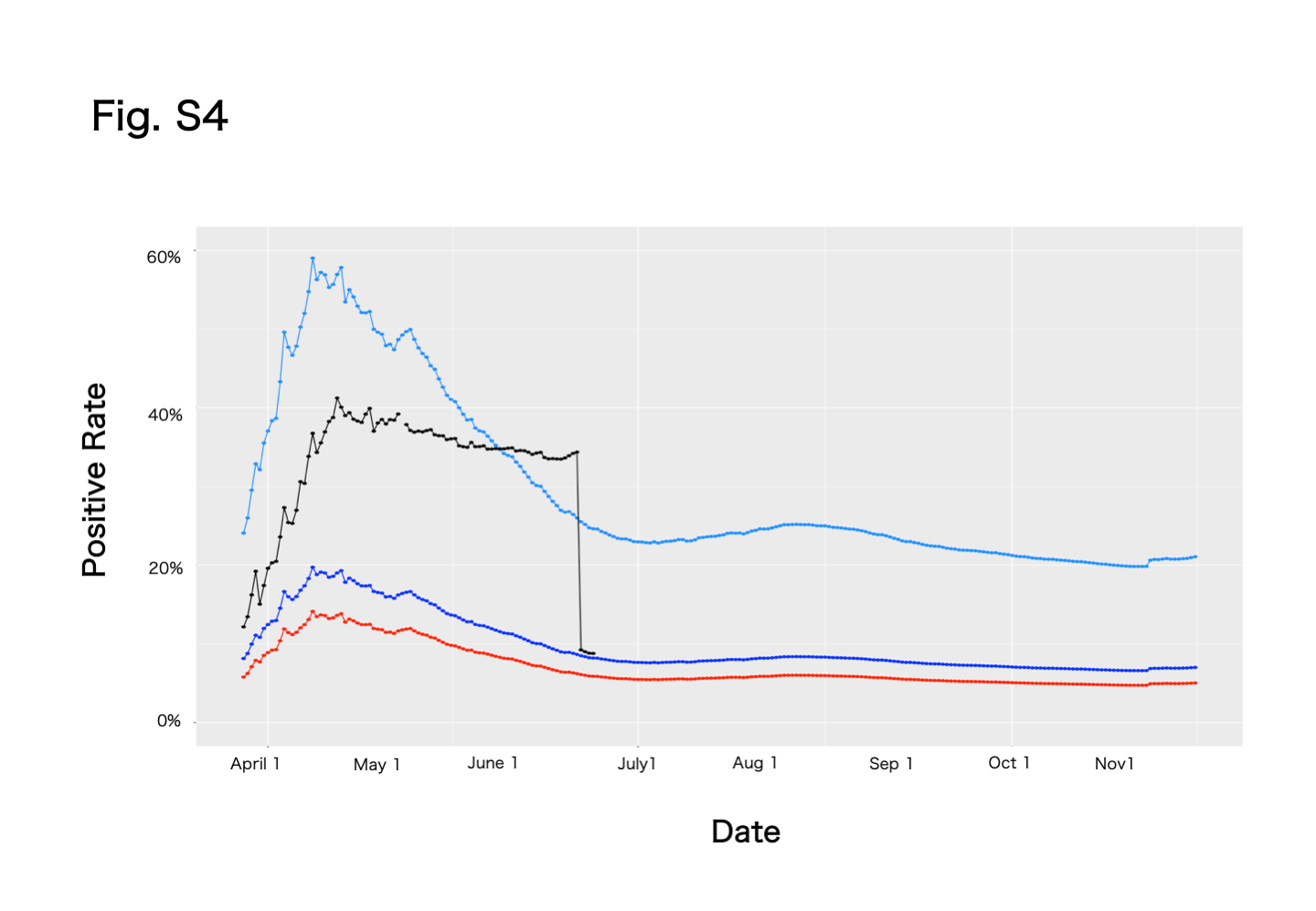
